# Admissions for eating disorders and other mental health diagnoses during the COVID-19 pandemic

**DOI:** 10.1101/2023.07.19.23292841

**Authors:** Sydney C. Jones, Megan Jacobs, Emile Latour, Rebecca Marshall, Michelle Noelck, Byron A. Foster

## Abstract

**Objective:** To examine how hospital admissions for mental health and eating disorders changed at the beginning of the COVID-19 pandemic and with the return to fully in-person school with increased vaccine availability.

**Methods:** Data from a tertiary care children’s hospital were examined for admissions to the hospital from March 2018 through March 2022, including children 6-20 years old admitted with ICD-10 codes for mental health and eating disorders. Interrupted time series (ITS) analyses were used to examine for changes at specific time points.

**Results:** In the time between March 2018 through March 2022, 851 admissions met inclusion criteria for eating disorders and 1,505 admissions for other mental health diagnoses. In the first year of the pandemic, the ITS analysis showed a significant increase in admissions per month for eating disorders with a slope of 1.2 (95% CI: 0.2, 2.2) and for other mental health diagnoses, a slope of 1.9 (95% CI: 1.1, 2.7). In a longer-term ITS analysis, return to fully in-person school was associated with a subsequent decrease in admissions over time at -1.0 per month (95% CI: - 2.1, 0.1). For other mental health diagnoses, return to school was associated with an initial drop and then an increase in admissions over time, slope of 2.2 (95% CI: -0.5, 3.8).

**Conclusion:** The COVID-19 pandemic had an initial impact on admissions for eating disorders and other mental health that attenuated over time. We note differences in the association of return to school on eating disorders compared with other mental health diagnoses.

## Introduction

The COVID-19 pandemic has drastically impacted the pediatric population. ^1^ When schools physically closed and students moved to primarily virtual learning, many children experienced social isolation from their peers and lost access to school-based mental health resources.^2^ Preliminary research suggests that mental health problems have increased among children and young adults since the onset of the COVID pandemic, with potential contributors including social isolation, difficulty accessing mental health resources, the economic recession, and the stress surrounding a public health crisis.^2,3,4,5^ There is emerging evidence from across the country that school closures had a detrimental impact on the health and wellbeing of adolescents. Two medical centers from the East Coast and Midwest found an increase in eating disorder hospital admissions in the months following their states’ stay at home orders.^6,7^ Other studies reported a general increase in eating disorder behavior since the start of the pandemic.^8,9,10,11,12,13^ Several hospitals reported increased emergency department mental health visits for pediatric patients during the later parts of the pandemic compared to before the pandemic.^14,15,16^

While several reports have described the initial increase in mental health-associated visits and admissions, few studies have described the impact of the overall trajectory of the COVID-19 pandemic,^1^ nor examined the association of changing pandemic school restrictions and vaccine availability on pediatric mental health hospital admissions. This study was conducted in Oregon where around 1500 to 1600 public, private, and charter schools tracked their virtual, hybrid, and in-person learning statuses. Following statewide school closures in March of 2020, by October 2020, approximately 83% of schools were using a virtual learning model. By the end of the 2020-2021 school year, only around 2% of the schools were virtual, with 61% of schools reporting hybrid instruction, and 34% reporting in person only.^17^ It was not until the start of the 2021-2022 school year, that all of Oregon schools returned to a mostly in-person full academic year.^18^ During this same time period, vaccine availability for the 12+ age group was approved for emergency use authorization (EUA) on May 12^th^ 2021.^19^ As of August 30, 2021, 52.1% of 12– 17-year-olds in Oregon had received one COVID-19 vaccine.^20^ The aim of this study was to examine how hospital admissions for mental health conditions and eating disorders changed at both the beginning of the pandemic and the start of the 2021-2020 school year, representing the first fully in-person school year and a greater return to pre-pandemic normalcy. We also examined whether there were differences in sex, gender, and racial and ethnic categories among children admitted for eating disorders and other mental health diagnoses over the course of the pandemic.

## Methods

### Study design and sample selection

A retrospective chart review of the electronic medical record was conducted at a 150-bed academic tertiary-care children’s hospital in the Pacific Northwest. We examined all admission encounters from March 1, 2018, two years before the onset of the COVID-19 pandemic, through March 31, 2022, for patients 6-20 years of age. Patients could be included more than once in the study period. Demographic data were abstracted from the medical record. We examined patients’ self-identified race and ethnicity as well as gender variables. Over the course of the study, the method used to categorize gender changed in the medical record, leading to a substantial number of patients with unspecified gender. We also examined biologic sex. This study was approved by our institutional IRB.

### Inclusion and exclusion criteria

Patients meeting the following criteria were included in this study: age 6-20 years and having an International Classification of Diseases, 10^th^ revision (ICD-10) code in their electronic health record for either the primary diagnosis or encounter diagnosis of an eating disorder or another mental health disorder including major depressive disorder, suicidal ideation, and intentional self-harm. Please see appendix for full list of ICD-10 codes.

We a priori defined children as having an eating disorder admission if they had an eating disorder ICD-10 code associated with their admission or an ICD-10 code of malnutrition that was not one of the excluded codes (e.g. Kwashiorkor). Similarly, we defined children as having a mental health admission if they had any mental health related ICD-10 codes associated with their admission encounter in the medical record (see appendix).

### Statistical analysis

Overall, we stratified the analysis by mental health diagnoses and eating disorder diagnoses. Descriptive statistics (means and standard deviations, medians and interquartile ranges, frequencies and percentages) were used to summarize patient characteristics. Group comparisons for continuous variables were made using one-way ANOVA, and comparisons for categorical variables using Fisher’s exact test. We conducted two interrupted time series (ITS) analyses: (1) examining the interruption of pandemic related school closures, and (2) examining the start of the first fully in person school year with vaccines available to adolescents.

The first ITS analysis examines the interruption of pandemic related school closures. The pandemic was assumed to start on April 1, 2020, just after the date of local pandemic-related school closures. The pre-pandemic period was considered as those hospital admissions between March 1, 2018 through March 31, 2020. The post-pandemic period includes the first year of pandemic admissions, April 1, 2020 through March 31, 2021.

The second ITS analysis examines whether the return to the fully in person academic year was associated with a change in hospital admissions. It examined two interruptions and three time periods: pre-pandemic period (March 1, 2018 to March 31, 2020); the pandemic school closure interruption (April 1, 2020); the pandemic year with school closure, hybrid school models, and some return to in-person learning (April 1, 2020 to August 31, 2021); the return to the fully in-person school year interruption (September 1, 2021); and the full school year return to in-person learning (September 1, 2021 to March 31, 2022). All analyses used R: A Language and Environment for Statistical Computing.^21^ A *p*-value < 0.05 was defined as statistically significant.

## Results

### All hospital admissions

All admitted patients from March 1, 2018 through March 31, 2022 can be seen in Figure 1. Before the pandemic, there were an average of 261.7 admissions per month (95% CI: 248.6, 274.8) with a stable slope over time of -0.5 admissions per month (95% CI: -1.7, 0.7; p=0.44). Starting April 1, 2020 there was a decrease in all admissions by -60.3 admissions (95% CI: -27.2, -93.4; p<0.001). Subsequently, the rate of admissions per month increased during the pandemic by 4.5 admissions per month (95% CI: 0.4, 8.5; p<0.001). On September 1, 2021, with the return to in-person learning, the number of admissions did not substantially change (p=0.60), and there was no change in the slope of admissions per month after that period (p=0.24) (Figure 1b). In summary, for all hospitalizations, we observed a decrease in admission numbers at the start of the pandemic, and then a slow rise in admission rate throughout the course of the pandemic lockdown and virtual to hybrid school period with no change in admission rate associated with the return to in person learning.

**Figure 1.**
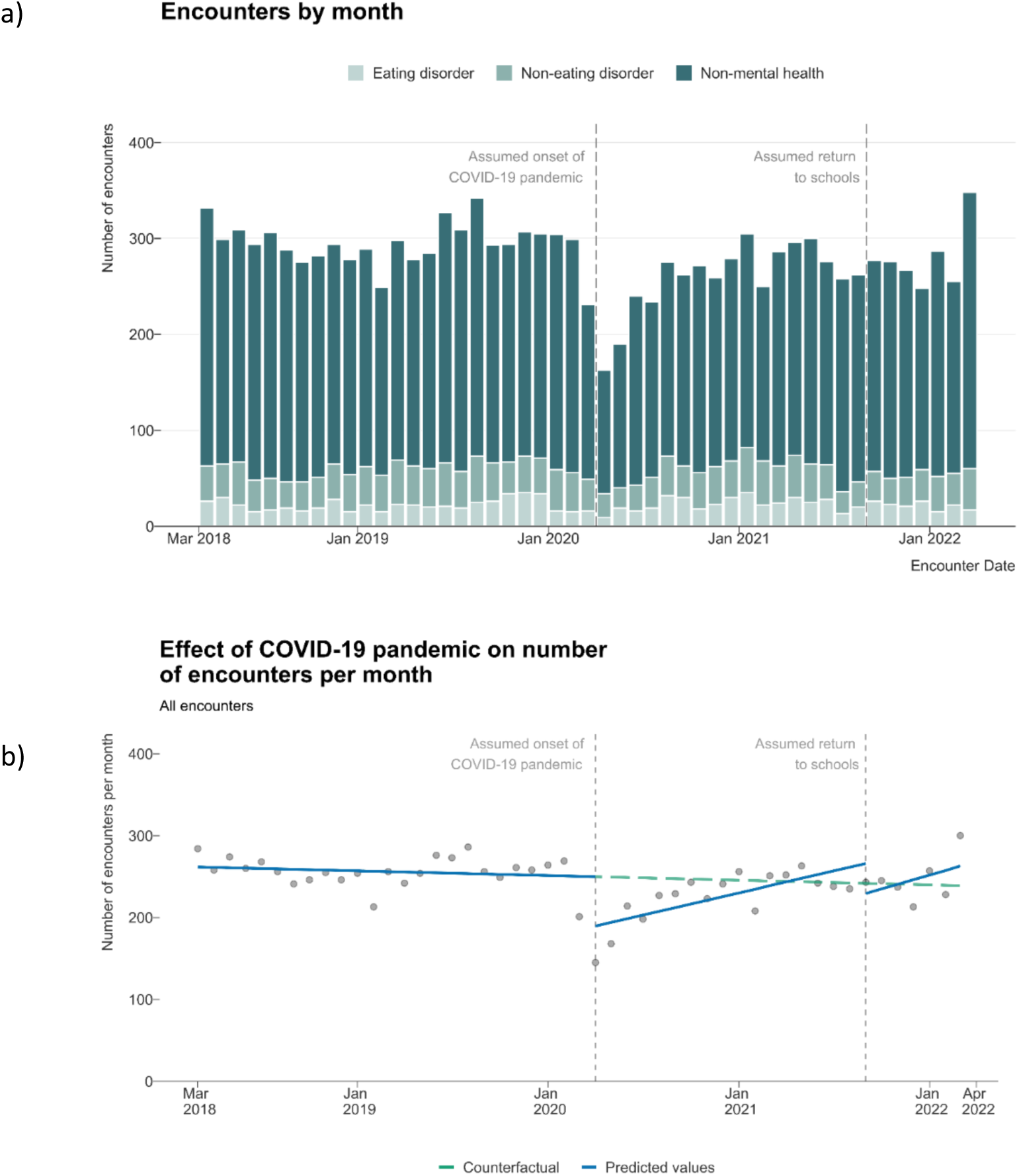
a) Graphical representation looking at hospital admissions by eating disorder, other mental health and non-mental health encounters over time and b) Interrupted time series analysis of the same data with onset of the pandemic and return to in-person school examined as interruptions

### Patient Characteristics

Between March 1, 2018 and March 31, 2022, there were a total of 851 patients who were admitted with a primary or secondary ICD-10 code for an eating disorder. During this same time period, there were 1,506 patients admitted with a mental health primary or secondary ICD-10 code. Patient characteristics can be found in Table 1. There was a relative increase in youth admitted with female sex for eating disorders after the onset of the pandemic (Table 1). An increase in the proportion of admissions for patients with a transgender or other gender was noted in the first part of the pandemic, increasing from 2.6% to 3.3% from April 1, 2020 through August 31, 2021, and then back down to 2.4% in the final period after return to school. For other mental health diagnoses, we saw a similar increase from 2.6% pre-pandemic to 4.2% during the April 1, 2020 through August 31, 2021 time frame, and a subsequent decrease to 1.6% post September 1, 2021. The proportion of those reporting Hispanic ethnicity decreased in both groups over time, as missing ethnicity data increased.

**Table 1.**
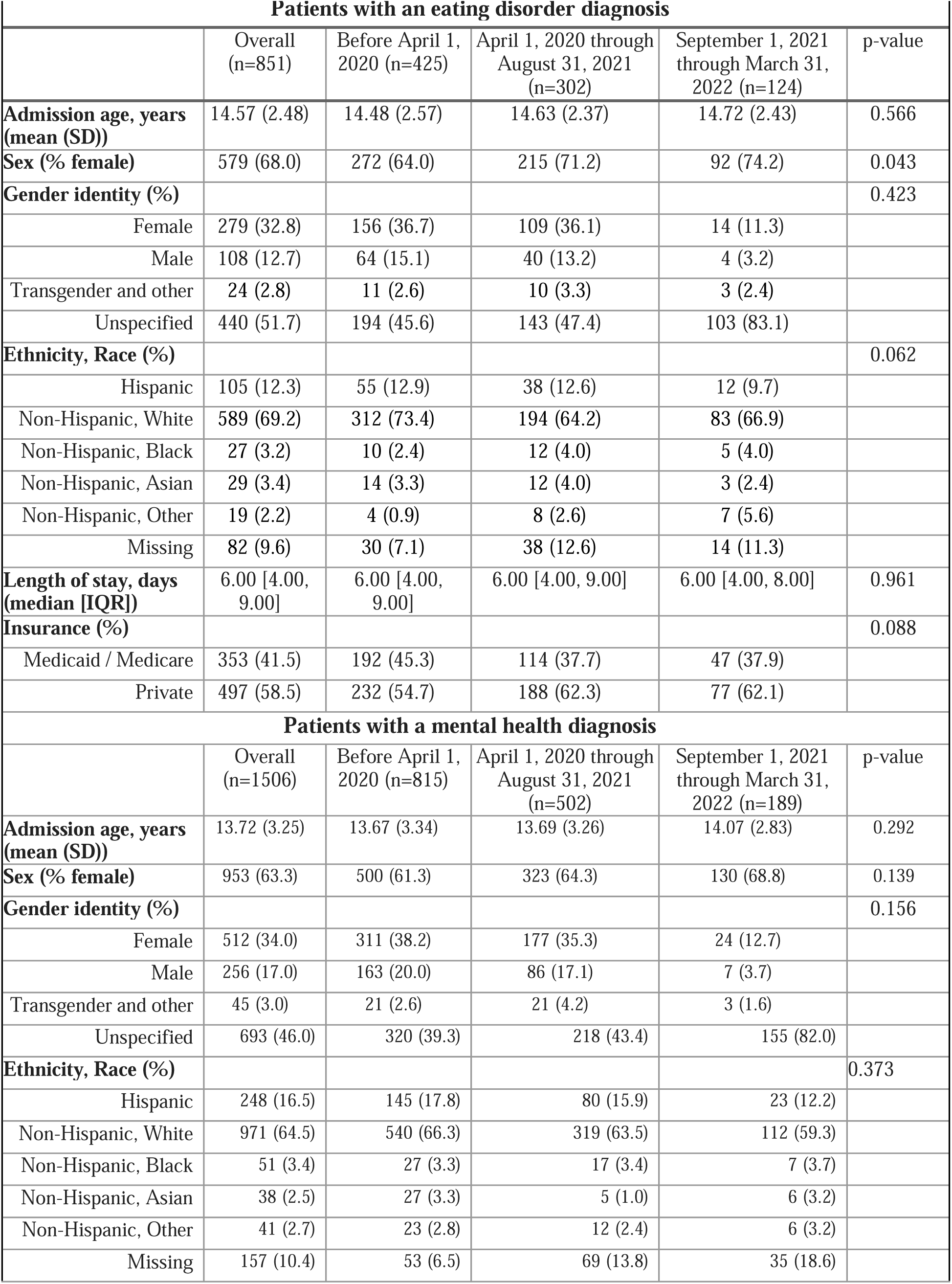
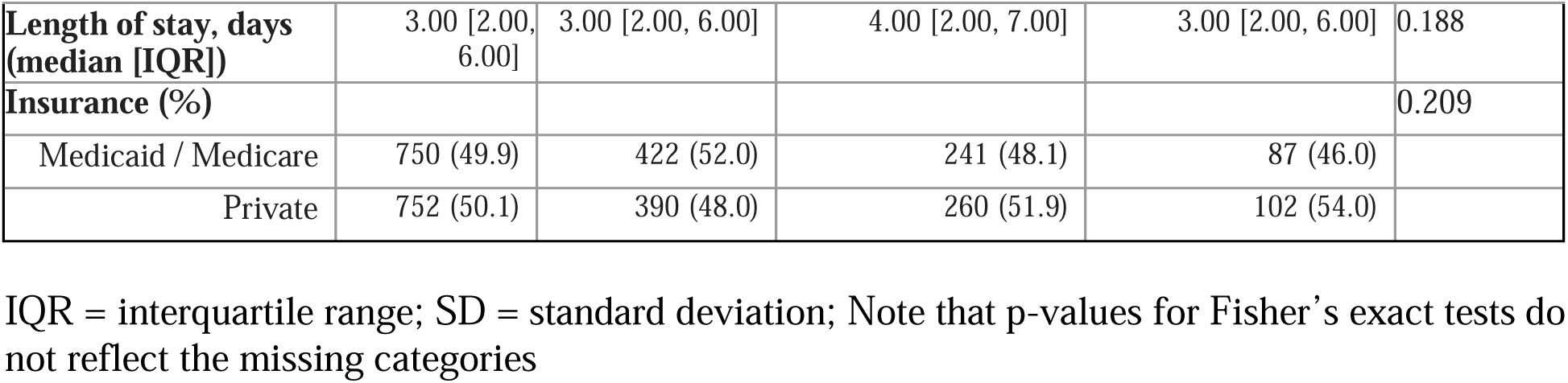
Characteristics of patients with admissions in the three time periods examined.

### Statistical Model

In the ITS analysis examining the effect of the pandemic interruption on April 1, 2020 and the subsequent year of admissions data, we modeled admissions separately for eating disorders (Figure 2) and other mental health diagnoses (Figure 3). The figures show the observed trend from the ITS models and the predicted trend had the pre-pandemic trends continued.

**Figure 2.**
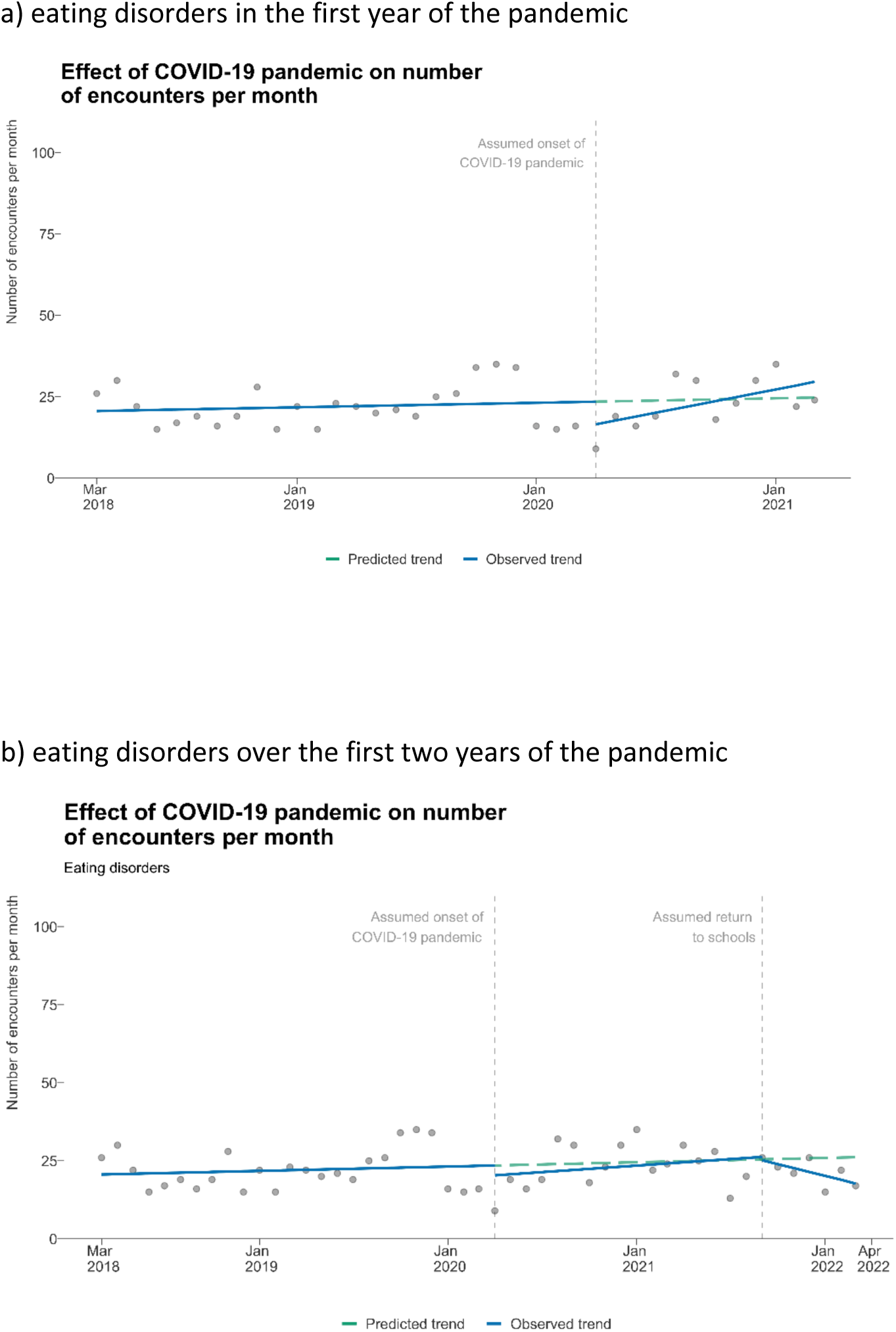
Interrupted time series analysis of admissions to the hospital with a diagnosis of eating disorder in a) the first year post-COVID and b) over the longer term (two years)

**Figure 3.**
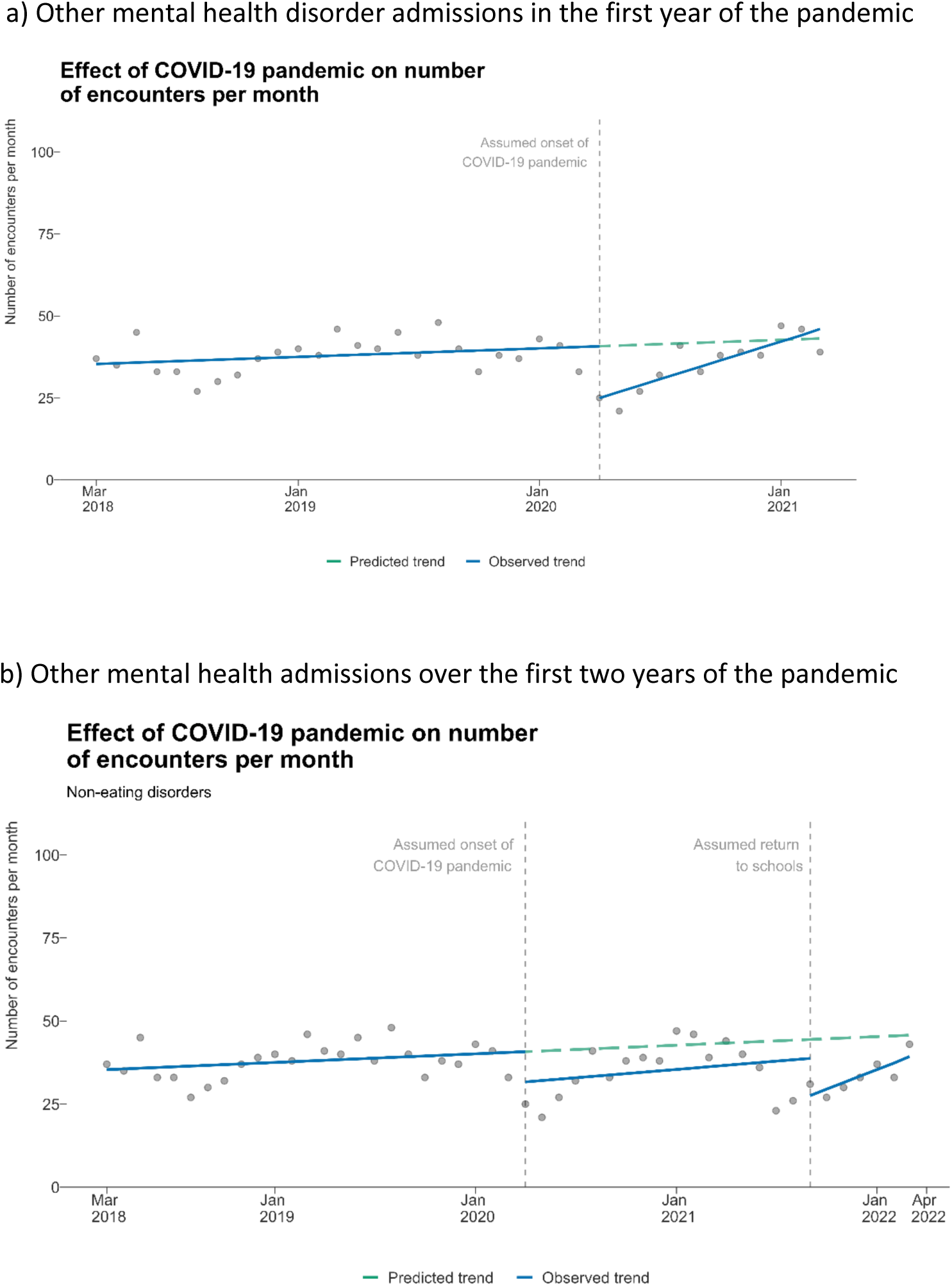
Interrupted time series analysis of admissions to the hospital with a diagnosis of other mental health (non-eating disorder) in a) the first year post-COVID and b) over the longer term (two years)

### Eating disorder admissions

For eating disorders, prior to the COVID-19 pandemic (prior to April 1, 2020), the average number of admissions per month was estimated to be 20.6 (95% CI: 15.9, 25.4). The rate of admissions per month was stable over time with a slope of 0.1 admissions per month (95% CI: -0.3, 0.5; p=0.588) (Figure 2).

On April 1, 2020, the number of eating disorder admissions showed a non-significant decrease of -7.0 (95% CI: -16.3, 2.3; p=0.142) admissions. The number of eating disorder admissions per month then increased over time during the first year of the pandemic with a slope of 1.2 (95% CI: 0.2, 2.2; p=0.020) admissions per month (Figure 2a).

The second set of ITS models (Figure 2b) included admissions over a longer time period and a second interruption on September 1, 2021, symbolizing time of increased socialization due to opening of more in-person school and increasing immunization rates. In examining the longer time period in this model, the number of admissions per month stayed the same during the pandemic with a slope of 0.3 admissions per month (95% CI: -0.4, 1.1; p=0.376). At the second interruption (September 1, 2021), the number of admissions remained stable at an estimated 0.3 admissions per month (95% CI: -14.2, 13.6; p=0.968). From September 1, 2021 through March 31, 2022, the number of admissions per month decreased slightly over time with a slope of -1.0 admissions per month (95% CI: -2.1, 0.1; p=0.072) without statistical significance (Figure 2b).

### Other mental health admissions

For other mental health diagnoses (Figure 3), prior to the COVID-19 pandemic, the average number of admissions per month was 35.4 (95% CI: 31.3, 39.5). The rate of admissions per month was stable over time with a slope of 0.2 admissions per month (95% CI: -0.1, 0.5; p=0.13).

At the first interruption on April 1, 2020, the number of mental health admissions decreased by 15.8 admissions (95% CI: 9.7, 21.9; p<0.001) (Figure 3a). Subsequently, the number of mental health admissions per month increased over time during the first year of the pandemic with a slope of 1.9 more admissions per month (95% CI: 1.1, 2.7; p<0.001).

Looking at the longer period of time of 18 months in the second ITS model (Figure 3b), the number of mental health admissions per month stayed the same over time during the pandemic with a slope of 0.4 admissions per month (95% CI: -0.5, 1.4; p=0.384). Using September 1, 2021 as the second interruption, the number of admissions decreased by -16.9 (95% CI: -26.5, -7.3; p<0.001). Subsequently, the number of admissions per month showed an increase with a slope of 2.2 admissions per month (95% CI: -0.5, 3.8; p=0.012).

## Discussion

The COVID-19 pandemic has had a significant impact on youth mental health.^1^ This study sought to determine impact at the level of hospitalizations at an urban tertiary care children’s hospital with a relationship to mental health etiologies including suicidality, depression, anxiety disorders, and medical stabilization for disordered eating. After the initial drop in admissions associated with the pandemic stay at home orders, the rate of both eating disorder and other mental health admissions significantly increased over the course of the first year of the pandemic, consistent with other reports.^6,7^ However, overall admissions over the longer course of the pandemic were stable compared to prior. Following the return to mostly in-person school with vaccines widely available in Oregon, we observed an initial decrease in mental health admissions suggesting that improved social interactions and possible perceived community safety related to increased immunization rates in youth may have acutely improved youth mental health. The significance in increased mental health admissions in the 2021 Fall and Winter months may reflect annual trends in depression, seasonal changes, or Omicron COVID-19 variant emergence in November of 2021.^22^ While these ecologic associations between major societal shifts and changes in admissions to hospitals have inherent epidemiological weaknesses in terms of causality, they can provide some insights into the effects associated with such changes.

The relative short-term increase in hospital admissions for disordered eating in the first year of the pandemic may be attributed to many factors, including the toll of pandemic stress and the changes in schooling on pediatric eating disorder health.^23,24^ Times of change or transition are associated with increases in and onset of eating disorders in adolescents.^25^ These findings are consistent with other studies demonstrating an increase in hospital admissions for eating disorders,^6,7,13^ increase in eating disorder behaviors, ^8,9,10,11^ and increase in eating disorder disease severity^12^ during the pandemic worldwide.

After lockdown, we observed a relative and short-term increase in admissions for both patients with eating disorders and other mental health diagnoses compared to the predicted hospitalization rate before the lockdown, similar to other interrupted time series on eating disorders hospital admissions before and during the pandemic.^6,7^ One possible explanation for this temporary increase in admissions includes the loss of school resources and structure for the pediatric patient population. It has been reported that of adolescents who use mental health services in a year, 57% used a school based mental health service and 35% accessed their mental health care exclusively from a school.^2^ Disruption of routines and peer connections, exposure to increased stress or abuse at home, and increased social media usage could also be contributors to increasing rates of admissions.^3^

Of interest is our finding that the return to school may have been associated with differential effects between eating disorders and other mental health admissions. While there are likely varied and multifactorial explanations for this finding, we hypothesize that the social isolation of school closure had variable impact on the two different patient populations. While there was lack of access to mental health resources through schools, it is possible that increased time spent with family units was in fact a protective factor for patients at risk for suicidality, in the midst of all of the societal stress of the pandemic.^26,27^ For adolescents with disordered eating, we hypothesize that more time spent in the home and with parent(s) could have led to higher awareness of the adolescents’ disordered eating behaviors that would have otherwise been easier to hide while at school. One other factor to consider with adolescents no longer being around peers in the school setting is the possibility that they experienced less bullying-which might have been a protective feature for all students.^27^ In addition, Oregon experienced one of the lowest COVID-19 infection rates per 100,000 citizens in the United States, and it is possible the stress of the pandemic did not affect Oregon’s pediatric psychiatric and eating disorder patient population as greatly as other states.^28,29^ There is a studied seasonality of pediatric mental health with increasing numbers of emergency department mental health presentations during the fall and spring and decreasing numbers during the summer.^30,31^ The hospital admissions greatly decreased in July with a slow increase towards September, consistent with the observed seasonality pattern of pediatric mental health. The level change also could be that after a complete lack of in-person school, returning to the classroom briefly improved pediatric mental health before the stresses of school contributed to increased hospital admissions.

We observed a slight increase in transgender patients admitted for both mental health and eating disorders in the pandemic period which may reflect increases in transgender identity among youth, which has been described recently using CDC data.^32^ However, this population also experiences discrimination and stress at baseline and may have been disproportionately impacted by the stressors of the pandemic. Given that only one third of transgender youth live in home with family acceptance,^33,34^ the extended school closures in Oregon may have been a particular challenge for these youth, in addition to increased barriers to transgender care.

Anecdotally, the severity of pediatric mental health and eating disorder admissions increased since April 2020. Need for complex care plans, psychiatry, psychology, and 1:1 observation safety needs were higher than normal. Changes to medical unit security and safety plans were reassessed frequently and dedicated nursing staff were called on to develop de-escalation plans to keep patients and staff protected. Those with disordered eating diagnoses were noted to have more significant bradycardia than had been experienced in the years prior in many cases, and patients reported more severe symptoms temporal to the onset of pandemic isolation (personal communication, medical director of acute care unit).

### Limitations

This study has some inherent limitations including examining ecological level changes and examining changes in admissions associated with those changes. Additionally, chart review of ICD-10 codes tied to a patient admission is complex. Codes are assigned based on many system and billing provider structures. For example, a patient being admitted is assigned a code, each consulting and primary billing provider assigns diagnostic codes to their visit each day, and there is a discharge diagnosis finally assigned. We were inclusive in examining the number of patients with an eating disorder, for example, by including all malnutrition diagnostic codes. This methodology may have included some patients with malnutrition without eating disorders.

## Conclusion

This study adds new information on the impact of the COVID-19 pandemic on children’s mental health using a period of two years of hospital admissions data post-pandemic start, highlighting both the initial increases observed in other studies but also importantly the association with return to in-person school. The COVID-19 pandemic highlighted many pre-existing challenges within our healthcare system— caring for patients with mental health crises and eating disorder diagnoses within a medical model of care is one of them.

## Supporting information

Supplemental Appendix ICD-10 Codes

## Data Availability

All data produced in the present work are contained in the manuscript.

## Acknowledgment

Jose Rodriguez extracted multiple iterations of the data from the medical records.

