## Supplemental Appendix ICD-10 Codes for "Admissions for eating disorders and other mental health diagnoses during the COVID-19 pandemic"

**Appendix of ICD-10 codes used for inclusion criteria in the study:**

The following codes were used and applied to the data extraction inclusive of all sub-codes within each category.

F40-F48: Anxiety, dissociative, stress-related, somatoform and other nonpsychotic mental disorders; F43: Reaction to severe stress, and adjustment disorders; F50: Eating disorders; F54: Psychological and behavioral factors associated with disorders or diseases classified elsewhere; F55: Abuse of non-psychoactive substances; F59: Unspecified behavioral syndromes associated with physiological disturbances and physical factors; F32: Major depressive disorder, single episode; F33: Major depressive disorder, recurrent; T14.91:Suicide attempt; R45.85 : Homicidal and suicidal ideations; X71-X83: Intentional self-harm; E40-E46 Malnutrition. Excluded codes were specific to Marasmus or Kwashiorkor types of malnutrition.
